# Neurobiological Underpinnings of Social-Emotional Impairments in Adolescents with Non-Suicidal Self-Injury

**DOI:** 10.1101/2020.11.12.20229138

**Authors:** Nina Lutz, Luca Villa, Nazia Jassim, Ian Goodyer, John Suckling, Paul Wilkinson

**Affiliations:** Department of Psychiatry, University of Cambridge, Cambridge, UK; Department of Psychiatry, School of Medicine, Yale University, New Haven, CT, USA; Department of Education and Psychology, Freie Universität Berlin, Berlin, Germany

**Keywords:** adolescent, facial expression, attentional bias, neurobiology, NSSI

## Abstract

**Objective:** Few studies have investigated the neurological underpinnings of social-emotional processing among individuals with non-suicidal self-injury (NSSI), despite the range of interpersonal impairments associated with the behavior. This study aims to identify NSSI-specific patterns of resting state functional connectivity (RSFC) and neural activation during an emotional facial expression task.

**Methods:** Participants were currently depressed, antidepressant-free adolescents with and without lifetime history of NSSI. Left and right amygdala were specified as seed regions for RSFC analysis (n=43 NSSI, n=9 clinical controls). The emotional faces task presented participants with neutral, happy, and sad faces. Whole-brain analyses examined neural activation during the task, and groups were compared on post-scan ratings of facial emotional intensity (n=39 NSSI, n=9 clinical controls).

**Results:** Groups did not differ in RSFC analyses. Adolescents with NSSI showed attenuated neural activation to happy (versus neutral) faces in areas of the occipital lobe and cerebellum, and rated neutral and sad faces as more negative than clinical controls.

**Conclusions:** While groups did not differ in baseline limbic connectivity, neurological and behavioral findings revealed NSSI-specific alterations in processing of social-emotional stimuli. Depressed adolescents with NSSI interpreted ambiguous or negative social stimuli more negatively than depressed controls, and had an attenuated neural response to positive social stimuli. This negative bias likely contributes to the myriad interpersonal difficulties associated with NSSI. Adolescents with NSSI may benefit from treatments which combat these negative social interpretations and improve control over emotional responses to interpersonal stress.

## Introduction

Approximately one in five adolescents (22%) engage in non-suicidal self-injury (NSSI), the intentional damage of body tissue without suicidal intent (Gillies et al., 2018). NSSI is most often used for intrapersonal reasons: to distract from unpleasant thoughts, decrease negative feelings, or generate positive sensations and emotions (Nock et al., 2009). However, NSSI can also serve interpersonal functions such as communicating distress, generating support, or fitting in with peers (Nock & Prinstein, 2005).

NSSI is associated with a range of social and emotional functional impairments (Turner et al., 2012). Individuals who engage in NSSI experience greater levels of negative emotions, emotional reactivity, emotion dysregulation (Anderson & Crowther, 2012; S. E. Victor & Klonsky, 2014), and alexithymia, a difficulty in identifying and describing emotions (Lüdtke et al., 2016). Adolescents with NSSI also report that, relative to their peers, they lack social skills (Claes et al., 2010), perceive social situations as more stressful (K. L. Kim et al., 2015), are more sensitive to social rejection (Brown et al., 2017; Perini et al., 2019), and select more maladaptive solutions to social problems (Nock & Mendes, 2008). These emotional and social impairments likely interact to contribute to NSSI engagement. Difficulties with social cue interpretation and deficits in interpersonal problem solving leave individuals prone to negatively interpreting social situations, and vulnerable to using NSSI to manage the resulting distress. In this way, interpersonal problems can act as a trigger for NSSI engagement (Hilt et al., 2008; Nock & Mendes, 2008). Indeed, research reveals that social rejection, interpersonal stress, and perceived criticism can precipitate NSSI engagement, and heightened sensitivity to social stress may contribute to the development and maintenance of NSSI (Miller et al., 2018; Nock et al., 2009; Tatnell et al., 2018).

The study of interpersonal functions and social processing is therefore highly relevant to understanding NSSI. However, the high prevalence of emotional impairments and limited emotional introspection in this population may render self-report measures unreliable. Researchers have therefore called for studies utilizing objective measures of social skills and emotion processing, including behavioral tasks and neuroimaging techniques (Bentley et al., 2014).

A growing body of fMRI literature supports a neurological basis for these deficits in samples with NSSI, including baseline alterations in resting-state connectivity of regions involved in social and emotional processing. Within the limbic system, the amygdala is a central structure in the processing of emotion and social cognition, including the interpretation of emotional facial expressions and initiation of emotional responses (Adolphs, 2006; Donegan et al., 2003; Haxby et al., 2002; Kesler-West et al., 2001). Additionally, the frontal regions are central to the regulation of emotional responses and impulsive decision making (Allman et al., 2001; Ghashghaei & Barbas, 2002; S. Kim & Lee, 2011). Impaired fronto-limbic connectivity would explain in part the impairments in decision making and heightened reactions to strong emotions underlying NSSI, as well as a tendency to perceive social stimuli negatively (Westlund Schreiner et al., 2015). Indeed, adolescents with NSSI exhibit disrupted amygdala RSFC compared to healthy controls (Westlund Schreiner et al., 2017), and aberrant amygdala connectivity is significantly associated with the frequency of NSSI engagement (Westlund Schreiner et al., 2018). However, several significant effects in these previous studies were eliminated when controlling for clinical symptoms. Comparing adolescents with NSSI to similar clinical controls without self-harm would improve our ability to accurately separate the neurological correlates of NSSI from those generally associated with psychopathology.

Facial emotion processing tasks are especially relevant in elucidating the neural mechanisms underlying impaired social-emotional functioning, as recognition and processing of emotional facial expressions is a critical early step in social interaction (Haxby et al., 2002; In-Albon et al., 2015; In-Albon, Bürli, et al., 2013). Amygdala hyperreactivity to facial expressions could lead to incorrect interpretation, predisposing individuals to intense and inappropriate emotional reactions to social interactions which are perceived negatively. There is some evidence to suggest that NSSI is associated with impairments in emotional facial expression recognition (Seymour et al., 2016), though results have been mixed (In-Albon et al., 2015).

Within the neurobiology literature, Westlund Schreiner and colleagues (Westlund Schreiner et al., 2017) found that adolescents with NSSI exhibit aberrant limbic activity during an emotional face processing task, including decreased amygdala-prefrontal cortex connectivity, compared to healthy controls. However, these significant results were eliminated when depressive symptom severity was included as a covariate, leading the authors to conclude that the observed amygdala-frontal circuit disturbances underlie both NSSI and the comorbid depressive symptoms reported by these adolescents - depression is associated with similar alterations in amygdala and prefrontal activation (Arnone et al., 2012; Cullen et al., 2014; Frodl et al., 2009; Phillips et al., 2003), as well as impairments in facial emotion recognition (Demenescu et al., 2010). Studies also associate NSSI with aberrant task-specific neural activation in paradigms involving social evaluation, rejection sensitivity, and social exclusion (Brown et al., 2017; Groschwitz et al., 2016; Malejko et al., 2019; Perini et al., 2019), suggesting NSSI-specific alterations of emotion regulation pathways in social contexts. However, many of these effects were only observed in comparisons to healthy controls, and studies did not control for clinical symptoms. It remains unclear whether NSSI is indeed related to altered neural response to social-emotional stimuli, beyond the confounding effects of comorbid psychopathology.

Additional research must therefore investigate neural correlates of NSSI within clinical samples of adolescents. The identification of neural circuitry associated with NSSI in populations at high risk for the behavior would allow for more effective evaluation and monitoring of NSSI risk, and contribute to the development of early interventions and preventative strategies for NSSI. Studies utilizing depressed controls are especially relevant; clinical samples reveal that at least two thirds of adolescents engaging in self-injury meet criteria for major depression (Glenn & Klonsky, 2013; In-Albon, Ruf, et al., 2013). In the present study, we investigate amygdala RSFC and whole brain neural activation during an emotional face processing task (happy, sad, and neutral faces) in a currently depressed clinical sample of adolescents, with or without a history of NSSI. Since antidepressant treatment is known to alter neural activation to happy and sad faces and reduce the negative processing bias associated with depression (Arnone et al., 2012; Sheline et al., 2001; T. A. Victor et al., 2013), we included only medication naïve adolescents.

We hypothesized that RSFC analyses would reveal abnormal fronto-limbic connectivity in the NSSI group, and abnormal connectivity with ROIs previously identified in the literature (Westlund Schreiner et al., 2017). In line with the negative social processing bias reported in other studies, we hypothesized that the NSSI group would show increased limbic activation while viewing sad faces and attenuated activation in response to happy faces. Since degree of neural activation correlates with the intensity of emotion expressed (Blair et al., 1999), we expected to see greater activity in response to high intensity versus low intensity sad faces. Finally, we expected the NSSI group to show a negative bias in their interpretation of facial emotions (i.e. rate facial expressions as more sad and less happy than clinical controls).

## Methods

### Participants

Participants were adolescents (aged 11-17 years) sampled from the IMPACT trial, a randomized controlled trial evaluating treatments of adolescent depression (Goodyer et al., 2017). The MR-IMPACT sub-study used neuroimaging to evaluate a subset of IMPACT trial participants from East Anglia and North London (Hagan et al., 2013). Inclusion criteria were current Major Depressive Disorder and a score of at least 27 on the Moods and Feelings Questionnaire (MFQ) (Costello & Angold, 1988) at initial assessment. Exclusion criteria included drug or alcohol dependence, learning disability, pregnancy or breastfeeding, brain abnormality, MRI contraindication, intolerance to the MRI environment, and use of medications which could adversely interact with SSRIs (Hagan et al., 2013). Participants and their parents gave written informed consent. Ethical approval for MR-IMPACT was provided by the Cambridgeshire 2 Research Ethics Committee (reference number 09/H0308/168), following the Declaration of Helsinki.

Participants for this study were sampled from MR-IMPACT based on the absence of current antidepressant medication use. The NSSI group reported lifetime history of NSSI, while clinical controls reported neither lifetime NSSI nor suicide attempts at baseline or during the 86-week IMPACT follow-up period. The final sample sizes were 43 adolescents with NSSI and 9 clinical controls for RSFC analysis, and 39 adolescents with NSSI and 9 clinical controls for task activation analysis. Demographic and clinical variables of the sample are presented in table 1.

**Table 1.** Group comparisons of demographic and clinical variables for RSFC and Emotional Faces Task analyses. Bold indicates significant group differences.

| RSFC | NSSI<br>*n* = 43 |  | Clinical Controls<br>*n* = 9 |  | *t* | *p* |
| --- | --- | --- | --- | --- | --- | --- |
|  | *M* | *SD* | *M* | *SD* |  |  |
| Age (years) | 15.50 | 1.16 | 15.52 | 1.32 | 0.055 | 0.96 |
| SMFQ score<sup>1</sup> | 17.65 | 5.41 | 15.67 | 3.87 | -1.04 | 0.30 |
| Number of diagnoses | 0.88 | 1.10 | 0.89 | 1.05 | 0.013 | 0.99 |
| | *n* | % | *n* | % | $\chi^2$ | *p* |
| Gender (female) | 37 | 86.05 | 6 | 66.67 | 1.95 | 0.16 |
| Ethnicity (white) | 38 | 88.37 | 6 | 66.67 | 2.69 | 0.10 |
| Anxiety diagnosis<sup>2</sup> | 14 | 32.56 | 4 | 44.44 | 0.46 | 0.50 |
| Behavioral diagnosis<sup>3</sup> | 8 | 16.60 | 1 | 11.11 | 0.29 | 0.59 |

| Emotional Faces Task | NSSI<br>*n* = 39 |  | Clinical Controls<br>*n* = 9 |  | *t* | *p* |
| --- | --- | --- | --- | --- | --- | --- |
|  | *M* | *SD* | *M* | *SD* |  |  |
| Age (years) | 15.69 | 1.11 | 15.26 | 1.20 | -1.03 | 0.31 |
| SMFQ score<sup>1</sup> | 18.15 | 5.65 | 14.33 | 4.58 | -1.88 | 0.066 |
| Number of diagnoses | 0.95 | 1.17 | 0.56 | 0.73 | -0.96 | 0.34 |
| | *n* | % | *n* | % | $\chi^2$ | *p* |
| Gender (female) | 35 | 89.74 | 5 | 55.56 | 6.15 | **0.013** |
| Ethnicity (white) | 36 | 92.31 | 6 | 66.67 | 4.40 | **0.036** |
| Anxiety diagnosis<sup>2</sup> | 15 | 38.46 | 3 | 33.33 | 0.082 | 0.78 |
| Behavioral diagnosis<sup>3</sup> | 7 | 17.95 | 1 | 11.11 | 0.25 | 0.62 |

<sup>1</sup> Short Moods and Feelings Questionnaire, measure of depressive symptoms in past two weeks
<sup>2</sup> Panic disorder, Agoraphobia, Separation Anxiety, Generalized Anxiety Disorder, Specific Phobia, Post Traumatic Stress Disorder
<sup>3</sup> Conduct Disorder, Oppositional Defiant Disorder

### Procedures

#### Psychopathology

Psychiatric diagnoses were assessed via the Kiddie Schedule for Affective Disorders, present and lifetime version (K-SADS-PL), a semi-structured interview using DSM-IV criteria (Kaufman et al., 1997). NSSI history was assessed via the Columbia-Suicide Severity Rating Scale (C-SSRS), a semi-structured clinical interview which assesses self-injurious thoughts and behaviors (Posner et al., 2011). During the interview, a researcher asks a series of questions about the nature of participants’ self-injury to determine suicidal intent. These include questions such as “Did you [hurt yourself] as a way to end your life? Or did you [hurt yourself] purely for other reasons, not at all to end your life or kill yourself (like to make yourself feel better, or get something else to happen)?”.

The Short Mood and Feelings Questionnaire (SMFQ) was administered at the time of scanning to assess depressive symptoms during the past two weeks (Angold et al., 1995). The SMFQ is derived from the longer Mood and Feelings Questionnaire (MFQ) (Costello & Angold, 1988), and has shown strong psychometric properties in child and adolescent samples (Angold et al., 1995; Sharp et al., 2006).

#### Emotional Faces Task

In an event-related fMRI paradigm, participants were presented with sad (low and high intensity), happy (low and high intensity), and neutral faces taken from the Facial Expressions of Emotion: Stimuli and Tests (Young et al., 2002). Participants were presented with each face for 2 seconds and then instructed to identify the gender of the actor by pressing a button. The task consisted of 96 randomized trials. Each trial began with a crosshair fixation and presented participants with one type of emotion at one level of intensity, interspersed with neutral faces to serve as a processing baseline.

After the fMRI scan, participants were again shown the images and instructed to rate the intensity of the emotion expressed (happy and sad) on a Likert scale from one (low intensity) to nine (high intensity). Additional task details have been previously published (Hagan et al., 2013).

#### fMRI data acquisition and processing

Scanning took place at the Wolfson Brain Imaging Centre, University of Cambridge, UK. A Siemens 3T Trim Trio scanner (Siemens, Surrey, England) was used to collect BOLD-sensitive echo-planar images (EPI) at a sampling time of 2.0 seconds. A quadrature birdcage headcoil was used to transmit and receive radio frequency pulses. An automatic shim was applied to maximize magnetic field homogeneity prior to data acquisition.

During resting-state data acquisition, 256 whole-brain images were collected over approximately nine minutes while participants were awake with eyes closed. During the task, 249 volumes of data were collected over approximately 8.5 minutes. Each volume of data was comprised of 32 slices 3.0 mm in thickness (TE = 30 ms, TR = 2 s, flip angle = 78°, and interleaved series), with the field of view 192 × 120 mm^2^ and voxel size 3.0 × 3.0 × 3.0 mm^3^.

Preprocessing included correction for head movement with FSL’s MCFLIRT (FMRIB’s Software Library, Motion Correction FMRIB’s Linear Image Registration Tool, www.fmrib.ox.ac.uk/fsl) (Jenkinson et al., 2002) using the speedypp algorithm from the BrainWavelet Toolbox (www.brainwavelet.org) according to published protocol (Patel et al., 2014). Fourier-space time-series phase-shifting was used to correct interleaved slice timing. Spatial smoothing was carried out at 6 mm. ANTs (Advanced Normalization Tools) (Avants et al., 2009) was used to non-linearly transform data to the standard Montreal Neurological Institute (MNI) space (Worsley, 2001) by FLIRT (FMRIB’s Linear Image Registration Tool). Images were overlaid with the Automated Anatomical Labelling (AAL) atlas (Tzourio-Mazoyer et al., 2002) with 116 defined regions of interest (ROIs). Five participants were excluded from RSFC analysis and nine were excluded from task activation analysis for excessive head motion, brain abnormalities, MRI artefacts, or pre-existing medical conditions. Additional details of data collection, processing, and analysis have been previously published (Chattopadhyay et al., 2017; Hagan et al., 2013; Patel et al., 2014).

### Data analysis

#### Statistical analysis

Analyses of behavioral, clinical, and demographic variables were conducted using STATA 12 (StataCorp., 2011). The significance threshold was set at p < 0.05. Linear regression were used to compare groups on task response times and post-scan ratings of facial emotional intensity based on the type of stimulus presented. Age, gender, and SMFQ scores were included as covariates. Demographic and clinical variable comparisons were conducted in a series of chi-squared tests and independent samples t-tests.

#### Resting State Functional Connectivity analysis

All analyses were conducted using FSL’s FLAME1 software (FMRIB’S Local Analyses of Mixed Effects, www.fmrib.ox.ac.uk). In line with previous research (Cullen et al., 2014; Westlund Schreiner et al., 2017, 2018), we carried out a seed-based whole-brain RSFC analysis with the left and right amygdala specified as seed regions. This was followed by a seed-based analysis using an ROI mask, again with the left and right amygdala specified as seed regions. ROIs were selected based on previous significant findings (Westlund Schreiner et al., 2017): prefrontal cortex, temporal lobe, occipital cortex, supplementary motor area, and anterior cingulate cortex. Analyses were done at the voxel level to determine whether any area within the ROIs was correlated with left or right amygdala activity.

Statistical analyses were performed using FSL’s FEAT (fMRI Expert Analysis Tool, www.fmrib.ox.ac.uk). Analyses employed a series of Generalized Linear Models (GLM) at each intracerebral voxel, relating the average time-series of each seed region (left and right amygdala) to the voxel time-series. Group-level GLMs (NSSI v. clinical controls) included demeaned age, gender, and depressive symptoms as covariates, based on past research relating these variables to alterations in amygdala RSFC and neural response during emotion processing (Cullen et al., 2014; Frodl et al., 2009; Wu et al., 2016). A Family-Wise Error (FWE) correction based on Gaussian Random Field Theory was used to correct for multiple testing with the cluster-forming voxel threshold specified at z >2.3 and p<0.05 (Worsley, 2001).

#### Task Activation Analysis

Whole-brain analyses were carried out to examine neural activation during task performance using FSL’s FEAT (www.fmrib.ox.ac.uk). First-level GLM analyses included five explanatory variables: i) neutral, ii) high intensity happy, iii) low intensity happy, iv) high intensity sad, and v) low intensity sad. Mixed effects group GLM analyses compared NSSI and clinical controls in four contrasts: i) happy versus neutral, ii) sad versus neutral, iii) high intensity happy versus low intensity happy, and iv) high intensity sad versus low intensity sad. Analyses included demeaned age, gender, and depressive symptoms as covariates, and FWE correction with the cluster-forming voxel threshold specified at z >2.3 and p<0.05 (Worsley, 2001).

## Results

### Descriptive Statistics

In the RSFC analysis, adolescents with NSSI (*n*=43) did not differ from clinical controls (*n*=9) on demographic or clinical variables. In the task activation analysis, adolescents with NSSI (*n*=39) were significantly more likely to be female (χ^2^(1, *n*=48)=6.15, *p*=0.013) and white (χ^2^(1, *n*=48)=4.40, *p*=0.036) than were clinical controls (*n*=9) (Table 1). Participants excluded from respective fMRI analyses did not significantly differ from those included on any demographic or clinical variable.

### Resting State Functional Connectivity

Participants with NSSI (n=43) did not differ from clinical controls (n=9) on amygdala RSFC in whole brain or ROI analyses.

### Emotional Faces Task Activation Analysis

The NSSI group (*n*=39) showed one cluster of significantly attenuated neural activation to happy versus neutral faces in areas of the occipital lobe and cerebellum, compared to clinical controls (*n*=9) (*p*=0.00011, FSL FEAT group-level GLM, FWE-corrected for multiple comparisons) (Table 2, Figure 2). Groups did not differ in neural response to sad versus neutral, high intensity happy versus low intensity happy, or high intensity sad versus low intensity sad faces.

**Table 2.** Whole-brain task activation analysis comparing NSSI (n=39) and clinical control (n=9) groups. Analyses included age, gender, and SMFQ score as covariates. One significant cluster emerged in the happy versus neutral face analysis.

| Contrast | Brain Regions | # of Voxels | MNI Coordinates of Peak Voxel (x, y, z) | Peak Z-value | Cluster $p$-value |
| --- | --- | --- | --- | --- | --- |
| Happy v. Neutral Faces | *Left* middle occipital gyrus, inferior occipital cortex, fusiform gyrus<br>*Right* crus I of cerebellar hemisphere<br>*Bilateral* calcarine sulcus, lingual gyrus, crus II of cerebellar hemisphere, lobule VI of cerebellar hemisphere<br>Lobule IV, V, VI, VII of vermis | 1647 | -12, -86, -26 | 3.53 | 0.00011 |

**Figure 2.**
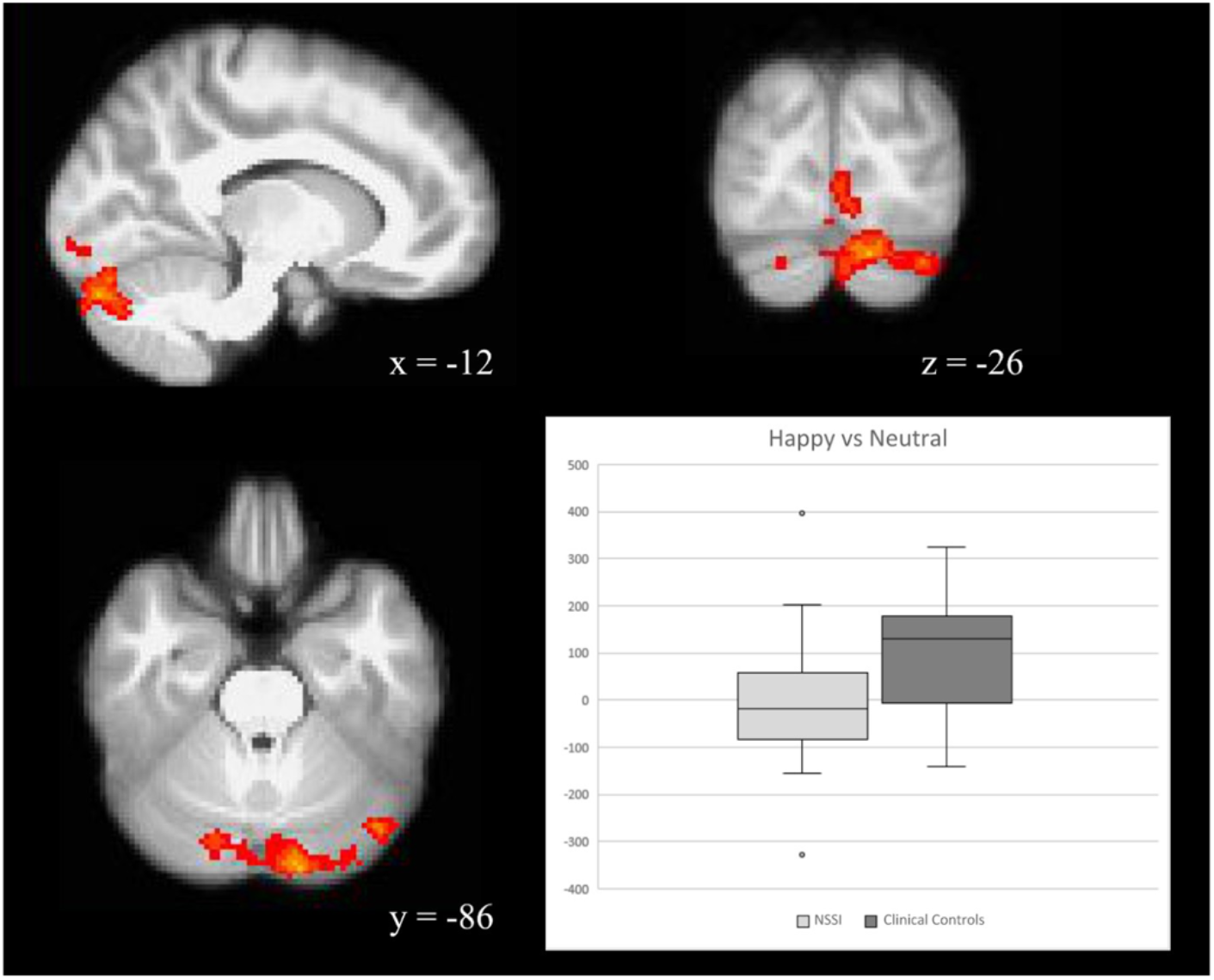
The NSSI group showed attenuated BOLD signal in the occipital lobe and cerebellum during happy versus neutral facial emotion processing. Coordinates are reported in MNI space. Box and whisker plot of BOLD signal illustrates the direction of the response.

### Emotional Faces Task Behavioral Performance

Groups did not significantly differ on mean reaction times based on valence or intensity of facial emotion. Groups significantly differed on post-scan rating of facial emotional intensity (Table 3); compared to clinical controls (*n*=9), the NSSI group (*n*=39) assigned significantly higher sad ratings to neutral faces (*F*(4,43)=1.51, *p*=0.033) and sad faces (*F*(4,43)=1.50, *p*=0.033), specifically low-intensity sad faces (*F*(4,43)=1.69, *p*=0.019).

**Table 3.**
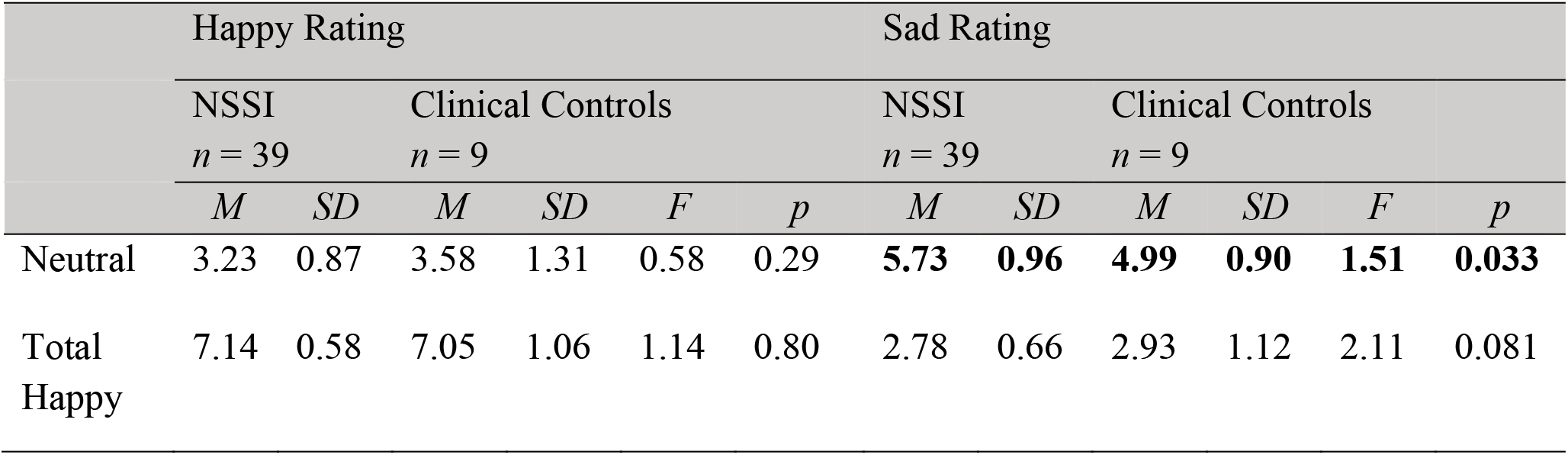
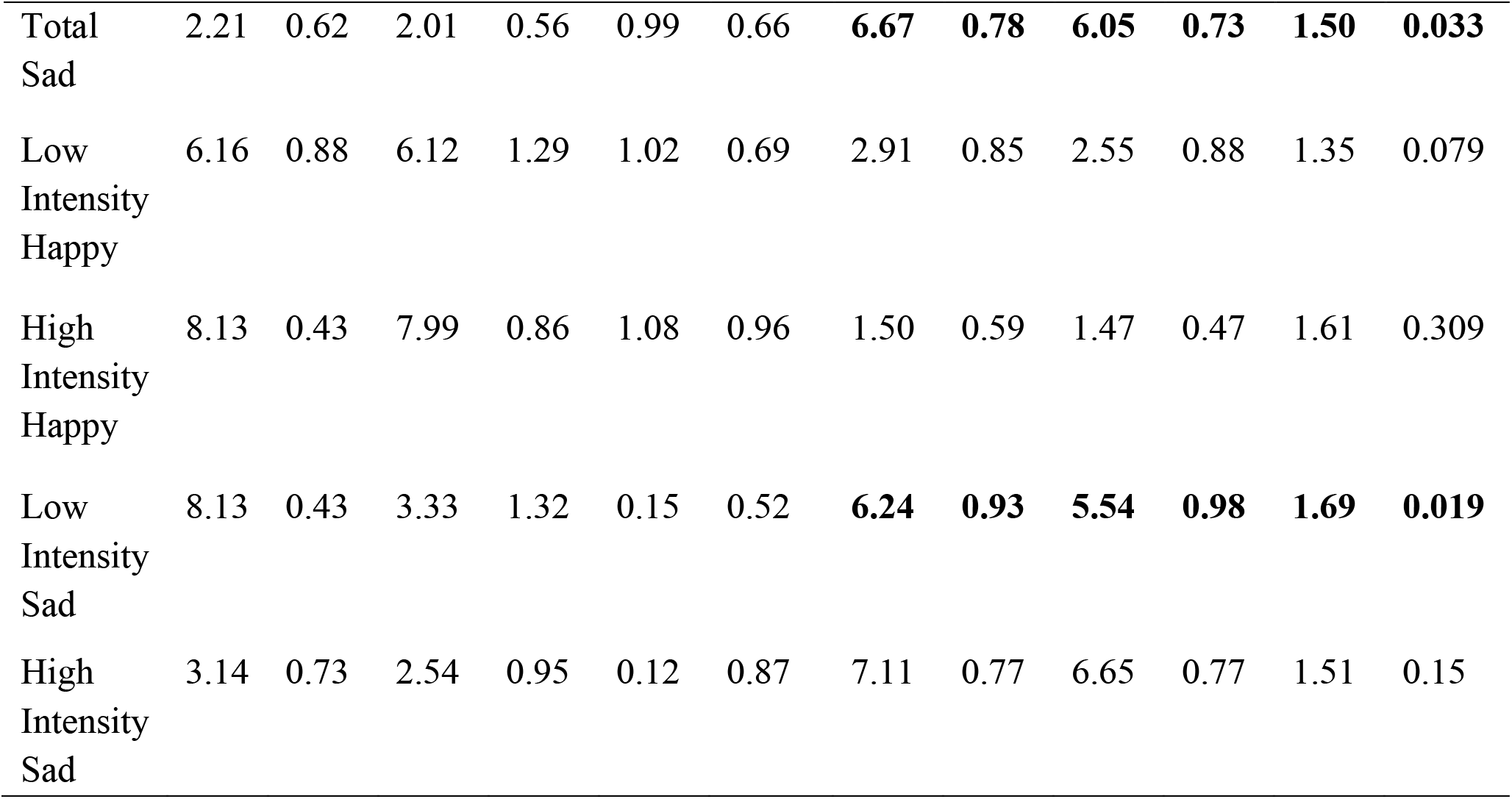
Emotional Faces Task, post-scan ratings of facial emotional intensity. Groups were compared in a series of linear regressions with age, gender, and SMFQ scores included as covariates. Bold indicates significance.

## Discussion

This study related NSSI to amygdala RSFC and neural activation during emotional facial expression processing in a clinical sample of currently depressed adolescents. In line with two previous publications (Westlund Schreiner et al., 2017, 2018), we expected to find abnormal limbic connectivity in the NSSI group. However, this hypothesis was not supported – we found no significant group differences in amygdala RSFC. Next, we hypothesized that adolescents with NSSI would show increased limbic activity specifically in response to sad faces, and expected to see an effect of emotional intensity. Surprisingly, groups did not differ in limbic activity during the task, and did not show any significant differences in contrasts of sad versus neutral or high intensity sad versus low intensity sad faces. This lack of positive findings was unexpected given the established role of negative affect in NSSI and the interpersonal impairments associated with the behavior. Only one significant cluster emerged: adolescents with NSSI exhibited diminished activity in regions of the occipital lobe and cerebellum during the processing of happy versus neutral faces.

The occipital lobe contains anatomical regions responsible for visual processing, and attenuated activation of the occipital lobe can signify reduced attention to visual stimuli (Li et al., 2013). Among healthy adolescents, processing of negative images elicits greater occipital cortex and fusiform gyrus activation, compared to neutral images (Stephanou et al., 2016). This suggests that emotional stimuli tend to elicit greater visual attention than neutral stimuli. In contrast, we found decreased activity in the occipital gyrus and fusiform gyrus, regions which are involved in the perception of face identity (i.e. recognition of a familiar face) (Haxby et al., 2002). This suggests an NSSI-specific reduction in attention to positive faces. This pattern of attention may contribute to a difficulty in picking up on social signals; adolescents with NSSI may not react as strongly to positive facial cues, and may therefore tend to perceive social situations more negatively than they actually are.

The NSSI group also showed reduced cerebellum activity in response to happy faces. The cerebellum is involved in a variety of social-emotional processes (Adamaszek et al., 2017; Van Overwalle et al., 2014), including the processing of emotional facial expressions (Baumann & Mattingley, 2012). We observed decreased activation in several regions which have been previously associated with specific emotion-related activity (E et al., 2014). Crus I and II are involved in the early steps of emotion perception and recognition, as well as the processing of emotional memory and the social expression of emotion (Adamaszek et al., 2017). Lobule VI plays a prominent role in emotion processing and displays functional connections to areas of the limbic system and the prefrontal cortex (Adamaszek et al., 2017). And finally, the vermis is involved in the process of forming, storing, and retrieving emotional memories. This area also interacts with the amygdala during emotion processing, and contributes to motor responses to emotion (Adamaszek et al., 2017). In line with findings of reduced visual attention in the occipital cortex, attenuated cerebellar activity suggests that adolescents with NSSI are impaired in the interpretation of and response to happy faces. NSSI may therefore be associated with difficulty in recognizing positive expressions in others, attributing these social cues to positive emotional memories, and having an appropriate emotional response. Furthermore, cerebellar dysfunction is related to impairments in social cognitive judgements, including “learning from the past and making strategic decision for the future” (Van Overwalle et al., 2014) (p.564). Aberrant cerebellum activity in response to social stimuli may therefore relate to impaired decision making in social contexts, which could leave individuals more prone to using NSSI as a means of social communication or in response to social triggers.

Finally, when comparing post-scan ratings of intensity of facial emotion expressed, the NSSI group assigned significantly higher sad ratings to neutral and sad faces, specifically low-intensity sad faces. This suggests that adolescents with NSSI are biased in their perception of potentially ambiguous facial expressions, tending to perceive them as negative. It is especially striking that this effect is evident in comparison to clinical controls, as depression is itself associated with negative perceptual biases (Naudin et al., 2014; Surguladze et al., 2004). A tendency to interpret neutral facial expressions as negative, and over-attribute negative emotion to sad expressions, could mean that adolescents with NSSI inaccurately infer that they are perceived negatively by others or that social interactions have gone poorly.

Taken together, our results do not support that NSSI is associated with aberrant limbic system activity or fronto-limbic connectivity, as researchers have suggested (Westlund Schreiner et al., 2017, 2018). This reinforces the importance of including clinical controls in NSSI studies. Several past neuroimaging studies compared individuals with NSSI to healthy controls, and may therefore have captured differences related to unspecified confounders – such as depression and borderline personality disorder symptoms (Donegan et al., 2003; Krause-Utz et al., 2014) – which are associated with self-injury, psychopathology, and aberrant limbic activity and connectivity.

Instead, we show that NSSI is associated with alterations in perception and response to socially ambiguous stimuli including neutral and low-intensity negative facial expressions, and deficits in the processing of positive facial expressions. The interpersonal disturbances associated with NSSI may be the result of biased perception of social-emotional stimuli, rather than an effect of emotional hyperreactivity. However, the observed differences in the occipital lobe and cerebellum were not pre-hypothesized, and so must be interpreted with caution.

### Limitations

A major strength of this study is the use of a clinical sample without current antidepressant medication use. However, a number of limitations affect the generalizability of our findings. First, we were limited by our small sample size and the lack of a healthy control group or a non-depressed NSSI group, since healthy controls in the IMPACT study were not assessed for NSSI. Second, we were limited by the NSSI data available on this sample; participants were only evaluated on a yes/no question about lifetime NSSI. Therefore, these adolescents may not have recent or repetitive NSSI, which has previously been associated with altered neural activity (Demers et al., 2019; Hooley et al., 2020; Westlund Schreiner et al., 2017, 2018). Third, the present task only included happy and sad faces, while previous significant findings have come in response to angry and fearful faces (Westlund Schreiner et al., 2017). Expressions of anger and fear in others could be perceived as a sign of social threat and therefore trigger limbic hyperreactivity among adolescents with NSSI. Happiness and sadness of others may hold less relevance to NSSI engagement.

### Future directions

Our findings suggest an additional neurally indexed perceptual disadvantage and attentional bias away from positive social stimuli in depressed adolescents with lifetime NSSI compared to those with depression alone. However, future research must investigate whether these neural differences do indeed relate to specific social-emotional functional impairments in samples with NSSI. Additional research is needed on the role of the cerebellum and occipital cortex in NSSI and in social-emotional processing - while past NSSI research has identified aberrant cerebellar and occipital activity at rest and in response to emotional stimuli (Plener et al., 2012; Westlund Schreiner et al., 2017), firm hypotheses on the role of these regions in NSSI have not been established. Longitudinal research should examine the interplay between NSSI and neurobiology. Research should continue to investigate the relationship between NSSI and the processing of social-emotional stimuli, and investigate treatments which improve social functioning and decrease the use of NSSI in response to social triggers.

## Data Availability

Data sharing is not applicable to this article as no new data were created or analyzed in this study.

## Acknowledgements

We sincerely thank the participants and their families, and the research assistants and clinicians who made the IMPACT and MR-IMPACT studies possible.

## References

Adamaszek, M., D’Agata, F., Ferrucci, R., Habas, C., Keulen, S., Kirkby, K. C., Leggio, M., Mariën, P., Molinari, M., Moulton, E., Orsi, L., Van Overwalle, F., Papadelis, C., Priori, A., Sacchetti, B., Schutter, D. J., Styliadis, C., & Verhoeven, J. (2017). Consensus Paper: Cerebellum and Emotion. Cerebellum, 16(2), 552–576. https://doi.org/10.1007/s12311-016-0815-8

Adolphs, R. (2006). Is the Human Amygdala Specialized for Processing Social Information? Annals of the New York Academy of Sciences, 985(1), 326–340. https://doi.org/10.1111/j.1749-6632.2003.tb07091.x

Allman, J. M., Hakeem, A., Erwin, J. M., Nimchinsky, E., & Hof, P. (2001). The anterior cingulate cortex. The evolution of an interface between emotion and cognition. Annals of the New York Academy of Sciences, 935(1), 107–117. https://doi.org/10.1111/j.1749-6632.2001.tb03476.x

Anderson, N. L., & Crowther, J. H. (2012). Using the experiential avoidance model of non-suicidal self-injury: Understanding who stops and who continues. Archives of Suicide Research, 16(2), 124–134. https://doi.org/10.1080/13811118.2012.667329

Angold, A., Costello, E. J., Messer, S. C., & Pickles, A. (1995). Development of a short questionnaire for use in epidemiological studies of depression in children and adolescents. International Journal of Methods in Psychiatric Research, 5(4), 237–249.

Arnone, D., McKie, S., Elliott, R., Thomas, E. J., Downey, D., Juhasz, G., Williams, S. R., Deakin, J. F. W., & Anderson, I. M. (2012). Increased amygdala responses to sad but not fearful faces in major depression: Relation to mood state and pharmacological treatment. American Journal of Psychiatry, 169(8), 841–850. https://doi.org/10.1176/appi.ajp.2012.11121774

Avants, B., Tustison, N., & Song, G. (2009). Advanced Normalization Tools (ANTS). Insight Journal. http://hdl.handle.net/10380/3113%0A

Baumann, O., & Mattingley, J. B. (2012). Functional topography of primary emotion processing in the human cerebellum. NeuroImage, 61(4), 805–811. https://doi.org/10.1016/j.neuroimage.2012.03.044

Bentley, K. H., Nock, M. K., & Barlow, D. H. (2014). The four-function model of nonsuicidal self-injury: Key directions for future research. Clinical Psychological Science, 2(5), 638–656. https://doi.org/10.1177/2167702613514563

Blair, R. J. R., Morris, J. S., Frith, C. D., Perrett, D. I., & Dolan, R. J. (1999). Dissociable neural responses to facial expressions of sadness and anger. Brain, 122(5), 883–893. https://doi.org/10.1093/brain/122.5.883

Brown, R. C., Plener, P. L., Groen, G., Neff, D., Bonenberger, M., & Abler, B. (2017). Differential neural processing of social exclusion and inclusion in adolescents with non-suicidal self-injury and young adults with borderline personality disorder. Frontiers in Psychiatry, 8, 267. https://doi.org/10.3389/fpsyt.2017.00267

Chattopadhyay, S., Tait, R., Simas, T., van Nieuwenhuizen, A., Hagan, C. C., Holt, R. J., Graham, J., Sahakian, B. J., Wilkinson, P. O., Goodyer, I. M., & Suckling, J. (2017). Cognitive Behavioral Therapy Lowers Elevated Functional Connectivity in Depressed Adolescents. EBioMedicine, 17, 216–222. https://doi.org/10.1016/j.ebiom.2017.02.010

Claes, L., Houben, A., Vandereycken, W., Bijttebier, P., & Muehlenkamp, J. (2010). Brief report: The association between non-suicidal self-injury, self-concept and acquaintance with self-injurious peers in a sample of adolescents. Journal of Adolescence, 33(5), 775– 778. https://doi.org/10.1016/j.adolescence.2009.10.012

Costello, E. J., & Angold, A. (1988). Scales to Assess Child and Adolescent Depression: Checklists, Screens, and Nets. Journal of the American Academy of Child and Adolescent Psychiatry, 27(6), 726–737. https://doi.org/10.1097/00004583-198811000-00011

Cullen, K. R., Westlund, M. K., Klimes-Dougan, B., Mueller, B. A., Houri, A., Eberly, L. E., & Lim, K. O. (2014). Abnormal amygdala resting-state functional connectivity in adolescent depression. JAMA Psychiatry, 71(10), 1138–1147. https://doi.org/10.1001/jamapsychiatry.2014.1087

Demenescu, L. R., Kortekaas, R., den Boer, J. A., & Aleman, A. (2010). Impaired attribution of emotion to facial expressions in anxiety and major depression. PLoS ONE, 5(12), e15058. https://doi.org/10.1371/journal.pone.0015058

Demers, L. A., Schreiner, M. W., Hunt, R. H., Mueller, B. A., Klimes-Dougan, B., Thomas, K. M., & Cullen, K. R. (2019). Alexithymia is associated with neural reactivity to masked emotional faces in adolescents who self-harm. Journal of Affective Disorders, 259, 253–261. https://doi.org/10.1016/j.jad.2019.02.038

Donegan, N. H., Sanislow, C. A., Blumberg, H. P., Fulbright, R. K., Lacadie, C., Skudlarski, P., Gore, J. C., Olson, I. R., McGlashan, T. H., & Wexler, B. E. (2003). Amygdala hyperreactivity in borderline personality disorder: Implications for emotional dysregulation. Biological Psychiatry, 54(11), 1284–1293. https://doi.org/10.1016/S0006-3223(03)00636-X

E, K. H., Chen, S. H. A., Ho, M. H. R., & Desmond, J. E. (2014). A meta-analysis of cerebellar contributions to higher cognition from PET and fMRI studies. Human Brain Mapping, 35(2), 593–615. https://doi.org/10.1002/hbm.22194

Frodl, T., Scheuerecker, J., Albrecht, J., Kleemann, A. M., Müller-Schunk, S., Koutsouleris, N., Möller, H. J., Brückmann, H., Wiesmann, M., & Meisenzahl, E. (2009). Neuronal correlates of emotional processing in patients with major depression. World Journal of Biological Psychiatry, 10(3), 202–208. https://doi.org/10.1080/15622970701624603

Ghashghaei, H. T., & Barbas, H. (2002). Pathways for emotion: Interactions of prefrontal and anterior temporal pathways in the amygdala of the rhesus monkey. Neuroscience, 115(4), 1261–1279. https://doi.org/10.1016/S0306-4522(02)00446-3

Glenn, C. R., & Klonsky, E. D. (2013). Nonsuicidal Self-Injury Disorder: An Empirical Investigation in Adolescent Psychiatric Patients. Journal of Clinical Child and Adolescent Psychology, 42(4), 496–507. https://doi.org/10.1080/15374416.2013.794699

Goodyer, I. M., Reynolds, S., Barrett, B., Byford, S., Dubicka, B., Hill, J., Holland, F., Kelvin, R., Midgley, N., Roberts, C., Senior, R., Target, M., Widmer, B., Wilkinson, P., & Fonagy, P. (2017). Cognitive behavioural therapy and short-term psychoanalytical psychotherapy versus a brief psychosocial intervention in adolescents with unipolar major depressive disorder (IMPACT): a multicentre, pragmatic, observer-blind, randomised controlled superiori. The Lancet Psychiatry, 4(2), 109–119. https://doi.org/10.1016/S2215-0366(16)30378-9

Groschwitz, R. C., Plener, P. L., Groen, G., Bonenberger, M., & Abler, B. (2016). Differential neural processing of social exclusion in adolescents with non-suicidal self-injury: An fMRI study. Psychiatry Research - Neuroimaging, 255, 43–49. https://doi.org/10.1016/j.pscychresns.2016.08.001

Hagan, C. C., Graham, J. M. E., Widmer, B., Holt, R. J., Ooi, C., van Nieuwenhuizen, A. O., Fonagy, P., Reynolds, S., Target, M., Kelvin, R., Wilkinson, P. O., Bullmore, E. T., Lennox, B. R., Sahakian, B. J., Goodyer, I., & Suckling, J. (2013). Magnetic resonance imaging of a randomized controlled trial investigating predictors of recovery following psychological treatment in adolescents with moderate to severe unipolar depression: Study protocol for Magnetic Resonance-Improving Mood with Psycho. BMC Psychiatry, 13, 247. https://doi.org/10.1186/1471-244X-13-247

Haxby, J. V., Hoffman, E. A., & Gobbini, M. I. (2002). Human neural systems for face recognition and social communication. Biological Psychiatry, 51(1), 59–67. https://doi.org/10.1016/S0006-3223(01)01330-0

Hilt, L. M., Cha, C. B., & Nolen-Hoeksema, S. (2008). Nonsuicidal Self-Injury in Young Adolescent Girls: Moderators of the Distress-Function Relationship. Journal of Consulting and Clinical Psychology, 76(1), 63–71. https://doi.org/10.1037/0022-006X.76.1.63

Hooley, J. M., Dahlgren, M. K., Best, S. G., Gonenc, A., & Gruber, S. A. (2020). Decreased Amygdalar Activation to NSSI-Stimuli in People Who Engage in NSSI: A Neuroimaging Pilot Study. Frontiers in Psychiatry, 11, 238. https://doi.org/10.3389/fpsyt.2020.00238

In-Albon, T., Bürli, M., Ruf, C., & Schmid, M. (2013). Non-suicidal self-injury and emotion regulation: a review on facial emotion recognition and facial mimicry. Child and Adolescent Psychiatry and Mental Health, 7(1), 5. https://doi.org/10.1186/1753-2000-7-5

In-Albon, T., Ruf, C., & Schmid, M. (2013). Proposed Diagnostic Criteria for the DSM-5 of Nonsuicidal Self-Injury in Female Adolescents: Diagnostic and Clinical Correlates. Psychiatry Journal, 2013, 159208. https://doi.org/10.1155/2013/159208

In-Albon, T., Ruf, C., & Schmid, M. (2015). Facial emotion recognition in adolescents with nonsuicidal self-injury. Psychiatry Research, 228(3), 332–339. https://doi.org/10.1016/j.psychres.2015.05.089

Jenkinson, M., Bannister, P., Brady, M., & Smith, S. (2002). Improved optimization for the robust and accurate linear registration and motion correction of brain images. NeuroImage, 17, 825–41. https://doi.org/10.1016/S1053-8119(02)91132-8

Kaufman, J., Birmaher, B., Brent, D., Rao, U., Flynn, C., Moreci, P., Williamson, D., & Ryan, N. (1997). Schedule for affective disorders and schizophrenia for school-age children-present and lifetime version (K-SADS-PL): Initial reliability and validity data. Journal of the American Academy of Child and Adolescent Psychiatry, 36(7), 980–988. https://doi.org/10.1097/00004583-199707000-00021

Kesler-West, M. L., Andersen, A. H., Smith, C. D., Avison, M. J., Davis, C. E., Kryscio, R. J., & Blonder, L. X. (2001). Neural substrates of facial emotion processing using fMRI. Cognitive Brain Research, 11(2), 213–226. https://doi.org/10.1016/S0926-6410(00)00073-2

Kim, K. L., Cushman, G. K., Weissman, A. B., Puzia, M. E., Wegbreit, E., Tone, E. B., Spirito, A., & Dickstein, D. P. (2015). Behavioral and emotional responses to interpersonal stress: A comparison of adolescents engaged in non-suicidal self-injury to adolescent suicide attempters. Psychiatry Research, 228(3), 899–906. https://doi.org/10.1016/j.psychres.2015.05.001

Kim, S., & Lee, D. (2011). Prefrontal cortex and impulsive decision making. Biological Psychiatry, 69(12), 1140–1146. https://doi.org/10.1016/j.biopsych.2010.07.005

Krause-Utz, A., Winter, D., Niedtfeld, I., & Schmahl, C. (2014). The Latest Neuroimaging Findings in Borderline Personality Disorder. Current Psychiatry Reports, 16(3), 438. https://doi.org/10.1007/s11920-014-0438-z

Li, J., Xu, C., Cao, X., Gao, Q., Wang, Y., Wang, Y., Peng, J., & Zhang, K. (2013).Abnormal activation of the occipital lobes during emotion picture processing in major depressive disorder patients. Neural Regeneration Research, 8(18), 1693--1701. https://doi.org/10.3969/j.issn.1673-5374.2013.18.007

Lüdtke, J., In-Albon, T., Michel, C., & Schmid, M. (2016). Predictors for DSM-5 nonsuicidal self-injury in female adolescent inpatients: The role of childhood maltreatment, alexithymia, and dissociation. Psychiatry Research, 239, 346–352. https://doi.org/10.1016/j.psychres.2016.02.026

Malejko, K., Neff, D., Brown, R. C., Plener, P. L., Bonenberger, M., Abler, B., & Graf, H. (2019). Neural Signatures of Social Inclusion in Borderline Personality Disorder Versus Non-suicidal Self-injury. Brain Topography, 32(5), 753–761. https://doi.org/10.1007/s10548-019-00712-0

Miller, A. B., Linthicum, K. P., Helms, S. W., Giletta, M., Rudolph, K. D., Hastings, P. D., Nock, M. K., & Prinstein, M. J. (2018). Reciprocal Associations Between Adolescent Girls’ Chronic Interpersonal Stress and Nonsuicidal Self-Injury: A Multi-wave Prospective Investigation. Journal of Adolescent Health, 63(6), 694–700. https://doi.org/10.1016/j.jadohealth.2018.06.033

Naudin, M., Carl, T., Surguladze, S., Guillen, C., Gaillard, P., Belzung, C., El-Hage, W., & Atanasova, B. (2014). Perceptive biases in major depressive episode. PLoS ONE, 9(2), e86832. https://doi.org/10.1371/journal.pone.0086832

Nock, M. K., & Mendes, W. B. (2008). Physiological Arousal, Distress Tolerance, and Social Problem-Solving Deficits Among Adolescent Self-Injurers. Journal of Consulting and Clinical Psychology, 76(1), 28–38. https://doi.org/10.1037/0022-006X.76.1.28

Nock, M. K., & Prinstein, M. J. (2005). Contextual features and behavioral functions of self-mutilation among adolescents. Journal of Abnormal Psychology, 114(1), 140–146. https://doi.org/10.1037/0021-843X.114.1.140

Nock, M. K., Prinstein, M. J., & Sterba, S. K. (2009). Revealing the Form and Function of Self-Injurious Thoughts and Behaviors: A Real-Time Ecological Assessment Study Among Adolescents and Young Adults. Journal of Abnormal Psychology, 118(4), 816– 827. https://doi.org/10.1037/a0016948

Patel, A. X., Kundu, P., Rubinov, M., Jones, P. S., Vértes, P. E., Ersche, K. D., Suckling, J., & Bullmore, E. T. (2014). A wavelet method for modeling and despiking motion artifacts from resting-state fMRI time series. NeuroImage, 95(100), 287–304. https://doi.org/10.1016/j.neuroimage.2014.03.012

Perini, I., Gustafsson, P. A., Hamilton, J. P., Kämpe, R., Mayo, L. M., Heilig, M., & Zetterqvist, M. (2019). Brain-based Classification of Negative Social Bias in Adolescents With Nonsuicidal Self-injury: Findings From Simulated Online Social Interaction. EClinicalMedicine, 13, 81–90. https://doi.org/10.1016/j.eclinm.2019.06.016

Phillips, M. L., Drevets, W. C., Rauch, S. L., & Lane, R. (2003). Neurobiology of emotion perception II: Implications for major psychiatric disorders. Biological Psychiatry, 54(5), 515–528. https://doi.org/10.1016/S0006-3223(03)00171-9

Plener, P. L., Bubalo, N., Fladung, A. K., Ludolph, A. G., & Lulé, D. (2012). Prone to excitement: Adolescent females with non-suicidal self-injury (NSSI) show altered cortical pattern to emotional and NSS-related material. Psychiatry Research - Neuroimaging, 203(2–3), 146–152. https://doi.org/10.1016/j.pscychresns.2011.12.012

Posner, K., Brown, G. K., Stanley, B., Brent, D. A., Yershova, K. V., Oquendo, M. A., Currier, G. W., Melvin, G. A., Greenhill, L., Shen, S., & Mann, J. J. (2011). The Columbia-suicide severity rating scale: Initial validity and internal consistency findings from three multisite studies with adolescents and adults. American Journal of Psychiatry, 168(12), 1266–1277. https://doi.org/10.1176/appi.ajp.2011.10111704

Seymour, K. E., Jones, R. N., Cushman, G. K., Galvan, T., Puzia, M. E., Kim, K. L., Spirito, A., & Dickstein, D. P. (2016). Emotional face recognition in adolescent suicide attempters and adolescents engaging in non-suicidal self-injury. European Child and Adolescent Psychiatry, 25(3), 247–259. https://doi.org/10.1007/s00787-015-0733-1

Sharp, C., Goodyer, I. M., & Croudace, T. J. (2006). The Short Mood and Feelings Questionnaire (SMFQ): A unidimensional item response theory and categorical data factor analysis of self-report ratings from a community sample of 7-through 11-year-old children. Journal of Abnormal Child Psychology, 34(3), 379–391. https://doi.org/10.1007/s10802-006-9027-x

Sheline, Y. I., Barch, D. M., Donnelly, J. M., Ollinger, J. M., Snyder, A. Z., & Mintun, M. A. (2001). Increased amygdala response to masked emotional faces in depressed subjects resolves with antidepressant treatment: An fMRI study. Biological Psychiatry, 50(9), 651–658. https://doi.org/10.1016/S0006-3223(01)01263-X

StataCorp. (2011). Stata Statistical Software: Release 12. In College Station, TX: StataCorp LP.

Stephanou, K., Davey, C. G., Kerestes, R., Whittle, S., Pujol, J., Yücel, M., Fornito, A., López-Solà, M., & Harrison, B. J. (2016). Brain functional correlates of emotion regulation across adolescence and young adulthood. Human Brain Mapping, 37(1), 7– 19. https://doi.org/10.1002/hbm.22905

Surguladze, S. A., Senior, C., Young, A. W., Brébion, G., Travis, M. J., & Phillips, M. L. (2004). Recognition Accuracy and Response Bias to Happy and Sad Facial Expressions in Patients with Major Depression. Neuropsychology, 18(2), 212–218. https://doi.org/10.1037/0894-4105.18.2.212

Tatnell, R., Hasking, P., Lipp, O. V., Boyes, M., & Dawkins, J. (2018). Emotional responding in NSSI: examinations of appraisals of positive and negative emotional stimuli, with and without acute stress. Cognition and Emotion, 32(6), 1304–1316. https://doi.org/10.1080/02699931.2017.1411785

Turner, B. J., Chapman, A. L., & Layden, B. K. (2012). Intrapersonal and interpersonal functions of non suicidal self-injury: Associations with emotional and social functioning. Suicide and Life-Threatening Behavior, 42(1), 36–55. https://doi.org/10.1111/j.1943-278X.2011.00069.x

Tzourio-Mazoyer, N., Landeau, B., Papathanassiou, D., Crivello, F., Etard, O., Delcroix, N., Mazoyer, B., & Joliot, M. (2002). Automated anatomical labeling of activations in SPM using a macroscopic anatomical parcellation of the MNI MRI single-subject brain. NeuroImage, 15(1), 273–289. https://doi.org/10.1006/nimg.2001.0978

Van Overwalle, F., Baetens, K., Mariën, P., & Vandekerckhove, M. (2014). Social cognition and the cerebellum: A meta-analysis of over 350 fMRI studies. NeuroImage, 86, 554– 572. https://doi.org/10.1016/j.neuroimage.2013.09.033

Victor, S. E., & Klonsky, E. D. (2014). Daily emotion in non-suicidal self-injury. Journal of Clinical Psychology, 70(4), 364–375. https://doi.org/10.1002/jclp.22037

Victor, T. A., Furey, M. L., Fromm, S. J., Öhman, A., & Drevets, W. C. (2013). Changes in the neural correlates of implicit emotional face processing during antidepressant treatment in major depressive disorder. International Journal of Neuropsychopharmacology, 16(10), 2195–2208. https://doi.org/10.1017/S146114571300062X

Westlund Schreiner, M., Klimes-Dougan, B., Begnel, E. D., & Cullen, K. R. (2015). Conceptualizing the neurobiology of non-suicidal self-injury from the perspective of the Research Domain Criteria Project. Neuroscience and Biobehavioral Reviews, 57, 381– 391. https://doi.org/10.1016/j.neubiorev.2015.09.011

Westlund Schreiner, M., Klimes-Dougan, B., & Cullen, K. (2018). F74. Frequency of Nonsuicidal Self-Injury Associated With Amygdala Functional Connectivity. Biological Psychiatry, 83(9 (supplement)), S266. https://doi.org/10.1016/j.biopsych.2018.02.687

Westlund Schreiner, M., Klimes-Dougan, B., Mueller, B. A., Eberly, L. E., Reigstad, K. M., Carstedt, P. A., Thomas, K. M., Hunt, R. H., Lim, K. O., & Cullen, K. R. (2017). Multi-modal neuroimaging of adolescents with non-suicidal self-injury: Amygdala functional connectivity. Journal of Affective Disorders, 221, 47–55. https://doi.org/10.1016/j.jad.2017.06.004

Worsley, K. (2001). Statistical analysis of activation images. In P. Jezzard, P. M. Matthews, & S. M. Smith (Eds.), Functional MRI: An Introduction to Methods (pp. 251–270). Oxford Medical Publications. https://doi.org/10.1093/acprof

Wu, M., Kujawa, A., Lu, L. H., Fitzgerald, D. A., Klumpp, H., Fitzgerald, K. D., Monk, C. S., & Phan, K. L. (2016). Age-related changes in amygdala-frontal connectivity during emotional face processing from childhood into young adulthood. Human Brain Mapping, 37(5), 1684–1695. https://doi.org/10.1002/hbm.23129

Young, a W., Perrett, D. I., Calder, a J., Sprengelmeyer, R., & Ekman, P. (2002). Facial expressions of emotion: stimuli and tests (FEEST). In Psychology. Thames Valley Test Company. https://doi.org/10.1016/S0010-0277(97)00003-6

